# Transcranial direct current stimulation-augmented cognitive training for post-COVID-19 cognition: A phase IIb randomized controlled trial

**DOI:** 10.64898/2026.05.11.26352906

**Authors:** Catalina Trujillo-Llano, Anna Elisabeth Fromm, Lydia Lingemann, Ulrike Grittner, Marcus Meinzer, Robert Fleischmann, Eva-Lotta Brakemeier, Daria Antonenko, Agnes Flöel

**Author notes:** Correspondence to: Prof. Dr. Agnes Flöel, Department of Neurology, University Medicine Greifswald, Fleischmannstraße 8, 17475 Greifswald, Germany.

## Abstract

**Background:** Cognitive dysfunction is a prevalent and debilitating symptom of post-COVID-19 condition with limited evidence-based interventions. Here, we assessed the efficacy of cognitive training (CT) alone and combined with transcranial direct current stimulation (tDCS) for cognitive enhancement in post-COVID-19 patients.

**Methods:** Neuromod-COV was a phase IIb, prospective, randomized, open-label, blinded-endpoint trial conducted at University Medicine Greifswald, Germany. The tDCS intervention was evaluated through a double-blind, sham-controlled design. Adults aged 18-60 with confirmed SARS-CoV-2 infection ≥ 6 weeks prior and post-infection cognitive complaints were eligible. Participants were randomly assigned (1:1:1) to CT with active tDCS (CT+AtDCS), CT with sham tDCS (CT+StDCS), or progressive muscle relaxation (PMR, non-cognitive control intervention) with sham tDCS. Intervention consisted of nine 20-minute sessions over three weeks of CT (letter updating task) or PMR with 2 mA tDCS (active/sham) applied over the left dorsolateral prefrontal cortex. The primary outcome was untrained working memory (WM; measured by N-back task accuracy) comparing CT with PMR at post-intervention. Secondary outcomes included trained and untrained WM, visuospatial memory, and self-report measures at post-intervention and 1-month follow-up comparing CT vs. PMR and CT+AtDCS vs. CT+StDCS. The trial was registered at ClinicalTrials.gov (NCT04944147).

**Results:** Between October 1, 2021, and August 7, 2024, 60 participants were randomized (76.7% female) to CT+AtDCS (*n* = 20), CT+StDCS (*n* = 20), or PMR (*n* = 20). CT did not improve untrained WM at post-intervention compared with PMR (primary outcome: *β* = 1.59, 95% CI - 1.30 to 4.48, *p* = 0.278; 1-back: *β* = 2.52, 95% CI -1.27 to 6.31, *p* = 0.191; 2-back: *β* = 0.66, 95% CI -3.12 to 4.44, *p* = 0.732). However, CT+AtDCS enhanced untrained WM at post-intervention and follow-up, and visuospatial memory at post-intervention compared with CT+StDCS (secondary outcomes). No intervention improved self-report outcomes. No serious adverse events occurred and incidence rate ratios were similar between groups.

**Conclusion:** CT alone did not improve untrained WM performance. However, CT with tDCS enhanced untrained WM and visuospatial memory, suggesting potential benefits of combined neuromodulation approaches for cognitive enhancement in post-COVID-19 patients.

## 1. Introduction

SARS-CoV-2 infection frequently results in persistent symptoms beyond the acute phase, affecting multiple organ systems and compromising physical, cognitive, and mental health domains [1]. Cognitive dysfunction is among the most prevalent post-acute sequelae, manifesting even after mild infections [2] and significantly impairing quality of life, daily functioning, and work capacity [3,4]. These cognitive deficits predominantly affect attention, memory, and executive function [5] and are associated with structural and functional brain alterations [6,7] alongside accelerated brain aging [8]. The underlying mechanisms likely involve neuroinflammation, blood-brain barrier disruption, microvascular injury, and hypoxia, which collectively contribute to neuronal damage and altered neurotransmission [9]. Evidence-based interventions for post-COVID-19 cognitive dysfunction remain scarce, with most treatment recommendations extrapolated from other conditions [10]. Effective and scalable treatments are urgently needed.

Cognitive training (CT) is a promising intervention for post-COVID-19 cognitive deficits [10]. Preliminary evidence suggests that self-administered CT through digital gaming interfaces can improve executive control and processing speed in affected patients [11]. However, CT typically enhances performance on trained tasks but demonstrates limited transfer to untrained cognitive tasks [12]. Transcranial direct current stimulation (tDCS), a non-invasive neuromodulation technique, may augment CT effects by modulating cortical excitability and promoting synaptic plasticity [13]. When combined with CT, tDCS facilitates both immediate and sustained transfer to untrained cognitive domains [14]. This combined approach has demonstrated efficacy in enhancing cognitive functions across clinical [15,16] and healthy aging populations [17]. Beyond cognitive improvements, this approach can induce structural and functional brain changes, including microstructural changes in gray and white matter and increased frontoparietal connectivity [18,19].

Previous tDCS studies in post-COVID-19 patients have shown improvements in fatigue, mood, and quality of life [20,21]. However, evidence for cognitive benefits remains limited and inconclusive. For example, a recent randomized controlled trial of CT with concurrent tDCS showed immediate improvements in inhibitory control, processing speed, and divided attention, but no effects on WM, episodic memory, or language [22]. Another randomized trial reported immediate cognitive improvements following CT with concurrent tDCS, but compared two active stimulation sites [motor cortex vs. dorsolateral prefrontal cortex (DLPFC)] without a sham-controlled condition, precluding conclusions about tDCS efficacy [23]. Other randomized controlled trials found no cognitive benefits from tDCS alone [20]. Similar null findings were observed when tDCS was combined with unstructured activities (e.g., reading, housework), in which dosage parameters were not standardized or monitored, limiting the interpretability of the findings [24]. To date, no randomized controlled trial has jointly evaluated the efficacy of CT and the additional benefits of tDCS augmentation on cognitive functions in post-COVID-19 patients.

We conducted Neuromod-COV, a phase IIb, prospective, randomized, open-label, blinded-endpoint (PROBE) trial with a novel three-arm design to address this gap. We evaluated the efficacy of CT compared with a control intervention (Progressive Muscle Relaxation; PMR) on cognitive, functional, and health-related quality of life measures in patients with post-COVID-19 cognitive symptoms. Participants with subjective cognitive complaints were enrolled to capture the clinical relevance of post-COVID-19 “brain fog” symptoms. To assess the additional benefit of tDCS, we included a double-blind, sham-controlled comparison within the CT groups, comparing CT with active tDCS (CT+AtDCS) to CT with sham tDCS (CT+StDCS). Treatment effects were evaluated on trained and untrained cognitive tasks, functional status, and health-related quality of life measures at post-intervention and 1-month follow-up.

## 2. Materials and methods

### 2.1. Study design

Neuromod-COV was a phase IIb, PROBE trial comparing CT with a control intervention (PMR). The secondary intervention, tDCS, was evaluated through a double-blind, sham-controlled comparison within CT groups (CT+AtDCS vs. CT+StDCS). The trial was conducted at the University Medicine Greifswald, Germany. The study was approved by the ethics committee of the University Medicine Greifswald (BB 066/21) and conducted in accordance with the Declaration of Helsinki. No patients or members of the public were involved in the design, conduct, or reporting of the trial. The study protocol was previously published [25]. The trial was registered at ClinicalTrials.gov (NCT04944147) and has been completed.

### 2.2. Participants

Eligible participants were adults aged 18-60 years with confirmed SARS-CoV-2 infection (PCR or antigen test) at least six weeks before study inclusion and subjective cognitive complaints following infection. Additional inclusion criteria included right-handedness and normal or corrected-to-normal vision. Exclusion criteria comprised acute COVID-19 illness, history of dementia or cognitive impairment before COVID-19, neurological or psychiatric disorders, psychotropic medication use, severe and untreated medical conditions, substance abuse history, and contraindication to tDCS. Participants were recruited from our local post-COVID outpatient clinic and regional community support groups. Participants did not receive any concomitant care or treatments targeting cognitive symptoms during the trial. Sex was self-reported, with “male” and “female” as available options. All participants provided written informed consent before study inclusion.

### 2.3. Randomization and masking

Participants were randomly assigned (1:1:1) to receive CT with active tDCS (CT+AtDCS), CT with sham tDCS (CT+StDCS), or PMR with sham tDCS. We used stratified block randomization with variable block sizes. Stratification was based on pre-intervention performance in the 2-back condition of the N-back task (accuracy ≤ 87% vs. > 87%) to ensure balanced cognitive performance across groups. The randomization sequence was generated using the *blockrand* package in R. An independent researcher, not involved in enrollment or outcome assessments, generated the randomization sequence and assigned participants to the groups. Allocation information was stored in a secure electronic spreadsheet and accessible only to the researcher responsible for randomization.

The behavioral interventions (CT and PMR) were open-label, while the tDCS intervention within CT groups (CT+AtDCS and CT+StDCS) was administered under double-blind conditions. Double-blinding was maintained using the stimulation device’s blinded mode, which prevented both participants and operators from identifying the stimulation condition. In sham conditions, a 30-second current was applied to mimic the initial sensation of active stimulation, then turned off. Outcome assessors and data analysts remained blinded to all group allocations throughout the study. Blinding success in the CT groups was evaluated at the final training session (visit 10) by asking participants to guess their stimulation condition with three response options: “active tDCS,” “sham tDCS,” or “don’t know.”

### 2.4. Procedures

Fig. 1 provides an overview of the study design. Participants completed nine 20-minute intervention sessions over three weeks, with three sessions per week on alternating days: Monday, Wednesday, and Friday. Each session involved either CT with active tDCS, CT with sham tDCS, or PMR with sham tDCS. The chosen training dose was based on previous CT and tDCS studies using the same protocol, which showed cognitive and neural benefits [18,19,26] while also maintaining high feasibility and adherence.

**Figure 1.**
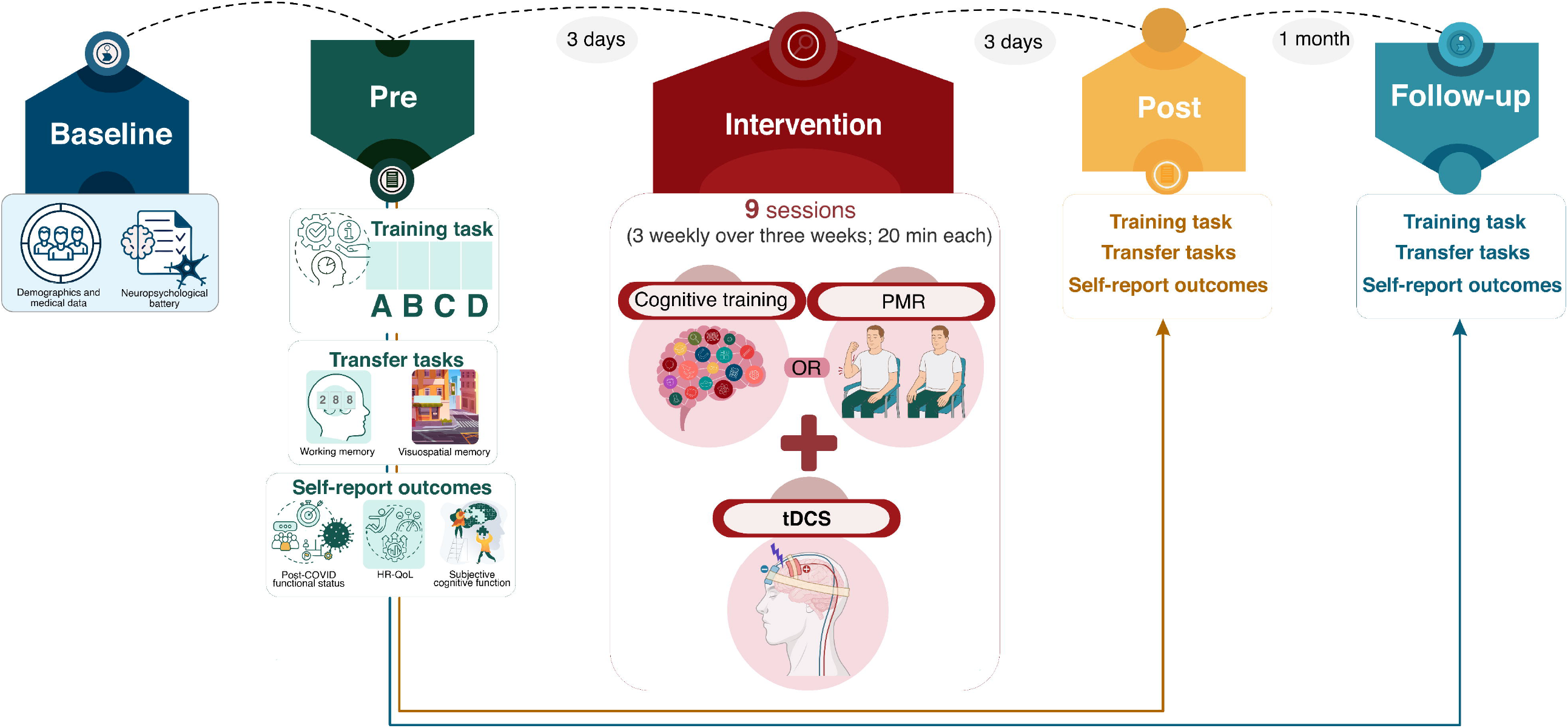
Study design. Participants were randomly assigned (1:1:1) to cognitive training with active tDCS (CT+AtDCS, *n* = 20), cognitive training with sham tDCS (CT+StDCS, *n* = 20), or progressive muscle relaxation with sham tDCS (PMR, *n* = 20). The intervention consisted of nine 20-minute sessions over three weeks (three sessions per week on alternating days). Baseline assessment included demographics, clinical, and cognitive characteristics. Outcome assessments were conducted at pre-intervention, post-intervention, and 1-month follow-up, including the training task (letter updating), transfer tasks (N-back for working memory; virtual reality maze for visuospatial memory), and self-report outcomes (Post-COVID-19 Functional Status Scale; PROPr Score for health-related quality of life; and PROMIS for subjective cognitive function). HR-QoL = health-related quality of life; PMR = progressive muscle relaxation; tDCS = transcranial direct current stimulation.

CT consisted of a tablet-based letter updating task targeting working memory (WM; [27]). Participants viewed lists of letters (A-D) of varying lengths (5, 7, 9, 11, 13, or 15 letters; six lists per length, totaling 36 lists) presented in random order, one letter at a time (2000 ms stimulus presentation; 500 ms inter-stimulus interval). After each list, participants were asked to recall the last four letters in the correct order. On the response screen, the four possible letters (A-D) appeared at the bottom, while four empty boxes appeared at the top. Participants placed the letters into the boxes in the correct order. Scores ranged from 0 to 36, with higher scores indicating better performance.

PMR followed Jacobson’s protocol [28]. A trained instructor guided participants through a standardized sequence of muscle tension (5-10 seconds) and relaxation (20-30 seconds), targeting major muscle groups and emphasizing the contrast between tension and relaxation. PMR served as a non-cognitive active control intervention, matching session duration, structure, and instructor contact while minimizing engagement of WM and executive processes targeted by the CT intervention. Although PMR can improve affective symptoms and sleep quality [29,30], direct cognitive-enhancing effects have not been demonstrated. PMR was delivered under sham tDCS to ensure comparable conditions to the CT intervention.

tDCS was administered concurrently with the CT or PMR interventions, with simultaneous onset. The stimulation was delivered using a battery-powered stimulator (Neuroelectrics Starstim 8, Barcelona, Spain) with a 4×1 montage targeting the left DLPFC (anode at F3, cathodes at F7, C5, FCz, AFz, according to the 10-20 EEG system). Electrodes (NG Pistim, 1 cm diameter) were positioned in a 64-channel neoprene headcap and filled with conductive saline gel (SignaGel). Active stimulation was delivered at 2 mA for 20 minutes, with 20-second ramp-up/down periods, maintaining stimulation throughout the intervention duration. Sham stimulation delivered 30 seconds of current to mimic initial sensations and then discontinued. A topical anesthetic (EMLA Cream, 25 mg/g Lidocaine + 25 mg/g Prilocaine) was applied to the electrode sites before stimulation to minimize skin sensations and enhance blinding.

Study assessments followed a standardized schedule. Baseline assessment (visit 0) included the collection of demographic, clinical, and neuropsychological data, as well as a brief practice session of the CT task. Outcome assessments were conducted at three timepoints: pre-intervention (visit 1, three days before the intervention started), post-intervention (visit 11, three days after the intervention was completed), and a 1-month follow-up (visit 12, one month after the post-intervention assessment). At each timepoint, participants completed two cognitive transfer tasks, the CT task and several self-report questionnaires (see below), which were administered in a fixed order across sessions. Parallel versions of the cognitive tasks were administered in a randomized counterbalanced order for each session.

### 2.5. Outcomes

#### 2.5.1. Primary outcome

The primary outcome was untrained WM performance comparing CT to PMR at post-intervention, measured by percentage accuracy on the N-back task. Participants viewed digits (1-9) presented sequentially (1500 ms stimulus presentation; 2500 ms inter-stimulus interval) and indicated via button press whether the current digit matched the one presented “n” positions earlier. The task consisted of 1-back and 2-back conditions, each with nine trials (10 stimuli per trial). Performance was calculated for each condition as percentage accuracy across all trials, with higher scores indicating better performance.

#### 2.5.2. Secondary outcomes

Secondary outcomes included cognitive afnd self-reported measures assessed at post-intervention and 1-month follow-up. Secondary cognitive outcomes included:

a. Near-transfer WM, assessed by N-back task accuracy (at follow-up for CT vs. PMR; at both timepoints for CT+AtDCS vs. CT+StDCS).
b. Far-transfer visuospatial memory, evaluated by the number of goals reached on the virtual reality maze task [31]. Participants encoded a predefined route and then recalled it by navigating to designated goal locations. The recall trial consisted of four 5-minute blocks during which participants could maximize the number of goals reached by correctly following the learned route as many times as possible within each block. Performance was quantified as the total number of goals reached across all blocks, with higher scores indicating better performance.
c. Trained WM, measured by the number of correctly recalled lists on the letter updating task [27].

Secondary self-reported outcomes included:

a. Functional status, assessed with the Post-COVID Functional Status Scale (PCFS; [32]), a 5-point ordinal rating of daily life limitations following COVID-19 (0 = no limitations, 1 = negligible limitations, 2 = slight limitations, 3 = moderate limitations, 4 = severe limitations).
b. Health-related quality of life, measured via the PROMIS-Preference (PROPr) score, [33] derived from the PROMIS-29 (Profile v2.1; [34]). Scores range from -0.022 to 1.0, with higher values indicating better overall health status.
c. Subjective cognitive function, evaluated using the PROMIS Cognitive Function Scale (Short Form 8a v2.0; [34]). Scores are expressed as standardized T-scores (mean = 50, SD = 10), with higher scores indicating better perceived cognitive function.

#### 2.5.3. Safety outcomes

Adverse events (AE) were assessed using a self-report questionnaire [35] administered at the end of every third training day (visits 4, 7, and 10). Participants rated the intensity of sensations, including itching, pain, burning, heat, metallic taste, fatigue, and other symptoms, on a 4-point scale (0 = none, 1 = mild, 2 = moderate, 3 = strong). For each participant, we calculated the number of moderate-to-strong sensations (scores > 1) across the three assessment points for each symptom category and the total number of events.

### 2.6. Statistical analysis

Sample size calculations are reported in the study protocol [25]. A total of 60 participants were required to detect an effect size of 0.8 (Cohen’s *d*) in the primary outcome, using an independent-samples *t*-test with 80% power and a two-tailed significance level of 0.05. All analyses followed a pre-registered statistical analysis plan (ClinicalTrials.gov: NCT04944147) and were conducted in the intention-to-treat population, defined as all randomized participants who received at least one intervention session. Missing data were estimated using multiple imputation by chained equations (30 imputed datasets, predictive mean matching). Per-protocol analyses, including only participants who completed all nine intervention sessions and scheduled visits, were performed as sensitivity analyses for all outcomes.

Primary and secondary outcomes were evaluated at post-intervention and 1-month follow-up. Two sets of group comparisons were conducted: (a) CT (pooled CT+AtDCS and CT+StDCS) vs. PMR and (b) CT+AtDCS vs. CT+StDCS. For continuous outcomes, linear mixed-effects models were fitted with post-intervention and follow-up scores as dependent variables. Fixed effects included group, timepoint, task condition (for the N-back task only), and their interactions. Pre-intervention scores, age, and sex were included as covariates, and random intercepts were included for the participants to account for autocorrelation of repeated measures. Between-group differences at each timepoint were estimated using model-based marginal means with 95% confidence intervals (CI).

Analyses comparing the CT groups used the same model structure, replacing the group variable with stimulation condition (active vs. sham). The only exception was the analysis of the CT task (letter updating) at post-intervention, in which all training sessions and post-intervention scores were included as the dependent variable. These models included the same fixed effects and covariates as above. Between-group differences at post-intervention were estimated using model-based marginal means with 95% CI.

For the analysis of functional status (PCFS) ordinal categories were collapsed due to sparse cell frequencies. Scores were dichotomized into two categories: negligible-to-slight (grades 1-2) and moderate-to-severe (grades 3-4). Binary logistic regression models with generalized estimating equations and a logit link function were fitted, using the same fixed effects and covariates as above. Group differences at each time point were estimated using model-based marginal probabilities as well as odds ratios with 95% CI.

Safety outcomes are reported as AE incidences (number of participants affected) and incidence rates with 95% CI) for the whole sample and by treatment group, based on the safety analysis set. Incidence rates (events per person-day) were calculated using Poisson regression models with the mean number of intervention days as an offset. Between-group comparisons were conducted using Poisson regression models with treatment group as the independent variable and the same offset term. Results are presented as incidence rate ratios with 95% CI.

Blinding success was assessed in the intention-to-treat population using the James Blinding Index (BI; [36]). The James BI ranges from 0 to 1, where 0 indicates complete unblinding, 0.5 indicates random guessing, and 1 indicates complete blinding. Unblinding was considered present if the upper limit of the two-sided 95% CI was below 0.5. All analyses were conducted in R (version 4.4.0). No interim analyses or stopping guidelines were planned or conducted.

## 3. Results

Between October 1, 2021, and August 7, 2024, 109 participants were screened for eligibility. Of these, 76 were invited to a baseline assessment, and 60 were randomly assigned to CT+AtDCS (*n* = 20), CT+StDCS (*n* = 20), or PMR (*n* = 20; Fig. 2). All participants received the allocated intervention, and all interventions were delivered as planned. All randomized participants were included in the intention-to-treat analysis. Four participants (6.7%; two in CT+AtDCS and two in CT+StDCS) discontinued the intervention due to logistical reasons. Eighteen participants (30%) had partial intervention completion: 14 (23.3%) missed one session, and four (6.7%) missed 2-5 sessions due to illness or logistical reasons, but continued with study assessments. The per-protocol analysis included 38 participants who completed the full intervention protocol and all assessments [CT+AtDCS (*n* = 12); CT+StDCS (*n* = 12); PMR (*n* = 14)].

**Figure 2.**
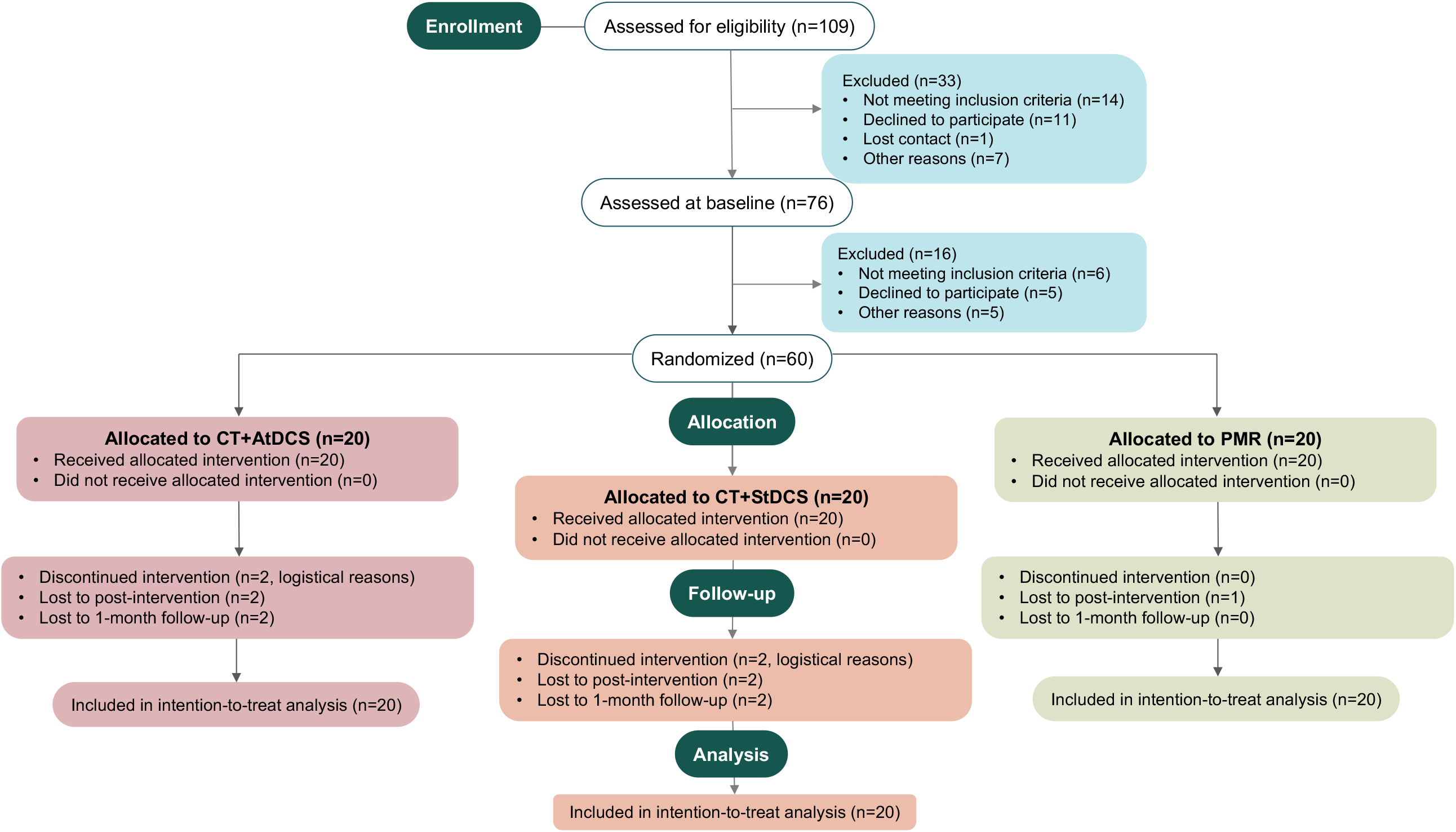
Trial profile. CT+AtDCS = cognitive training with active transcranial direct current stimulation; CT+StDCS = cognitive training with sham transcranial direct current stimulation; PMR = progressive muscle relaxation.

Baseline demographic, clinical, and cognitive characteristics were similar between groups (Table 1 and Supplementary Table S1). Participants had a mean age of 42.4 years (*SD* = 12.8), a mean of education of 16.2 years (*SD* = 3.5), and 76.7% were female. The mean time since COVID-19 infection was 13.4 months (*SD* = 8.7).

**Table 1.** Baseline demographics and clinical characteristics.

|  | Overall<br>(*n* = 60) | CT<br>(*n* = 40) | PMR<br>(*n* = 20) | CT+AtDCS<br>(*n* = 20) | CT+StDCS<br>(*n* = 20) |
| --- | --- | --- | --- | --- | --- |
| **Demographics** |  |  |  |  |  |
| Age (years) | 42.4 (12.8) | 43.8 (12.5) | 39.7 (13.2) | 43.5 (11.0) | 44.0 (14.2) |
| Education (years) | 16.2 (3.5) | 16.2 (3.6) | 16.1 (3.4) | 15.8 (3.3) | 16.6 (3.8) |
| Sex (female) | 46 (76.7%) | 30 (75.0%) | 16 (80.0%) | 17 (85.0%) | 13 (65.0%) |
| **Medical history** |  |  |  |  |  |
| Time since COVID infection (months) | 13.4 (8.7) | 12.2 (8.5) | 15.9 (8.8) | 12.2 (7.0) | 12.3 (10.0) |
| Hypertension | 8 (13.3%) | 7 (17.5%) | 1 (5.0%) | 1 (5.0%) | 6 (30.0%) |
| Diabetes | 3 (5.0%) | 2 (5.0%) | 1 (5.0%) | 2 (10.0%) | 0 (0.0%) |
| Cardiac arrhythmia | 5 (8.3%) | 3 (7.5%) | 2 (10.0%) | 0 (0.0%) | 3 (15.0%) |
| Kidney dysfunction | 1 (1.7%) | 0 (0.0%) | 1 (5.0%) | 0 (0.0%) | 0 (0.0%) |
| Liver dysfunction | 1 (1.7%) | 0 (0.0%) | 1 (5.0%) | 0 (0.0%) | 0 (0.0%) |
| Vitamin B12 deficiency | 2 (3.3%) | 1 (2.5%) | 1 (5.0%) | 0 (0.0%) | 1 (5.0%) |
| Hypothyroidism | 7 (11.7%) | 5 (12.5%) | 2 (10.0%) | 3 (15.0%) | 2 (10.0%) |
| Cardiovascular disease | 0 (0.0%) | 0 (0.0%) | 0 (0.0%) | 0 (0.0%) | 0 (0.0%) |
| **Other symptoms** |  |  |  |  |  |
| Fatigue | 56.4 (7.2) | 56.0 (8.0) | 57.2 (5.4) | 56.7 (6.8) | 55.3 (9.1) |
| Anxiety | 50.8 (6.6) | 51.1 (6.8) | 50.1 (6.3) | 52.0 (6.0) | 50.1 (7.5) |
| Depression | 49.9 (6.6) | 50.3 (6.6) | 48.9 (6.7) | 50.7 (5.9) | 50.0 (7.3) |
| Sleep disturbance | 51.5 (8.1) | 51.8 (8.9) | 50.8 (6.3) | 53.2 (7.5) | 50.4 (10.1) |

Data are mean (SD) or *n* (%). Other symptoms are PROMIS-29 Profile v2.1 standardized T-scores. CT = cognitive training; CT+AtDCS = cognitive training with active transcranial direct current stimulation; CT+StDCS = cognitive training with sham transcranial direct current stimulation; PMR = progressive muscle relaxation.

Results for primary and secondary outcomes in the intention-to-treat population are presented in Fig. 3, along with the corresponding raw scores in Fig. 4 and Supplementary Tables S2 and S3. Results for primary and secondary outcomes in the per-protocol population are presented in Supplementary Fig. S1.

**Figure 3.**
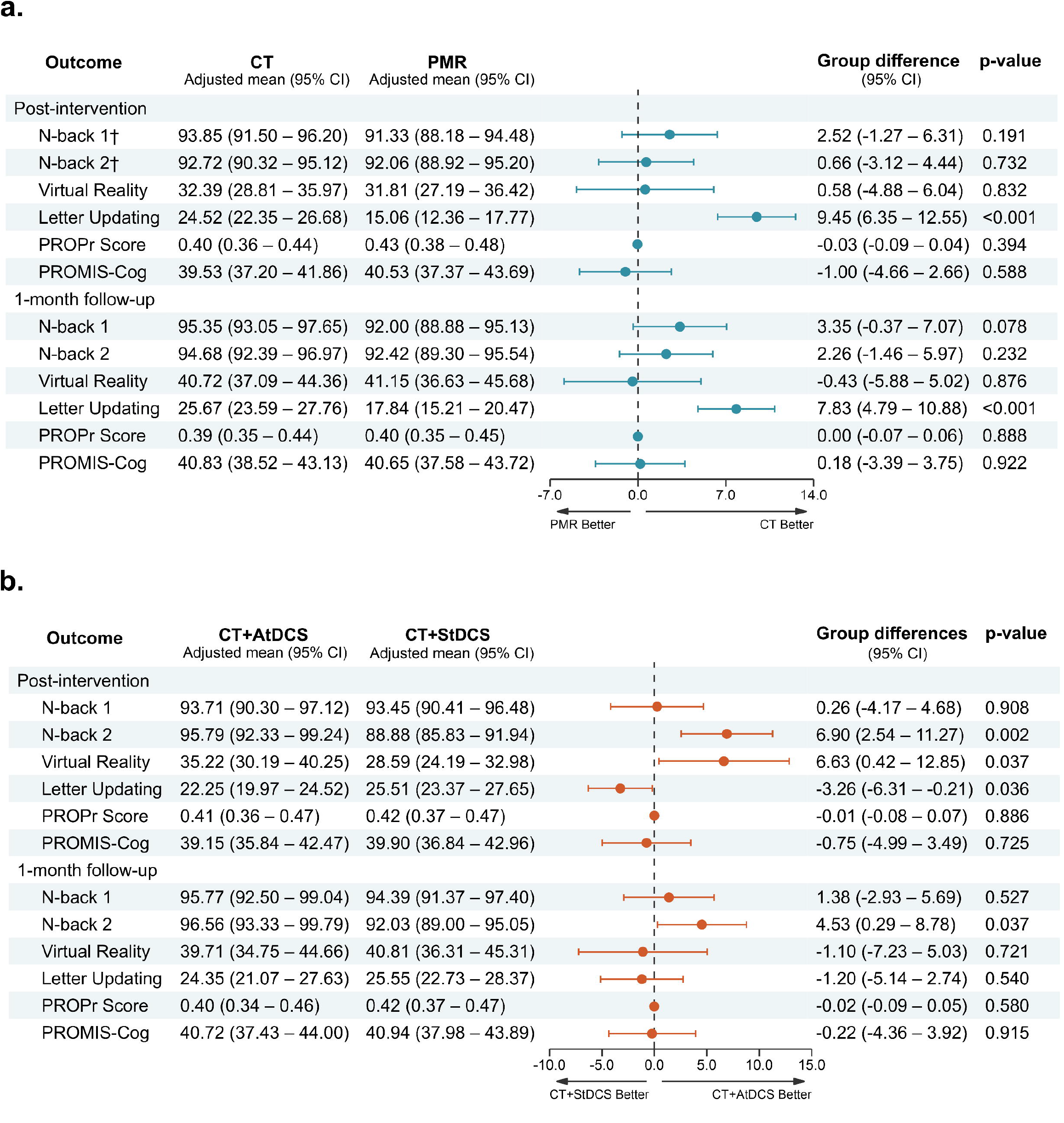
Results for primary and secondary outcomes in the intention-to-treat population. **(a)**Cognitive training (*n* = 40) vs. progressive muscle relaxation (*n* = 20). **(b)** Cognitive training with active tDCS (n = 20) vs. cognitive training with sham tDCS (*n* = 20). Forest plots show adjusted between-group mean differences with 95% CI at post-intervention and 1-month follow-up, derived from linear mixed-effects models using multiple imputation. Positive values favor cognitive training in panel (a) and cognitive training with active tDCS in panel (b). ^†^Primary outcome. CT = cognitive training; CT+AtDCS = cognitive training with active transcranial direct current stimulation; CT+StDCS = cognitive training with sham transcranial direct current stimulation; PMR = progressive muscle relaxation; PROMIS-Cog = PROMIS Cognitive Function Scale; PROPr = PROMIS-Preference Score.

**Figure 4.**
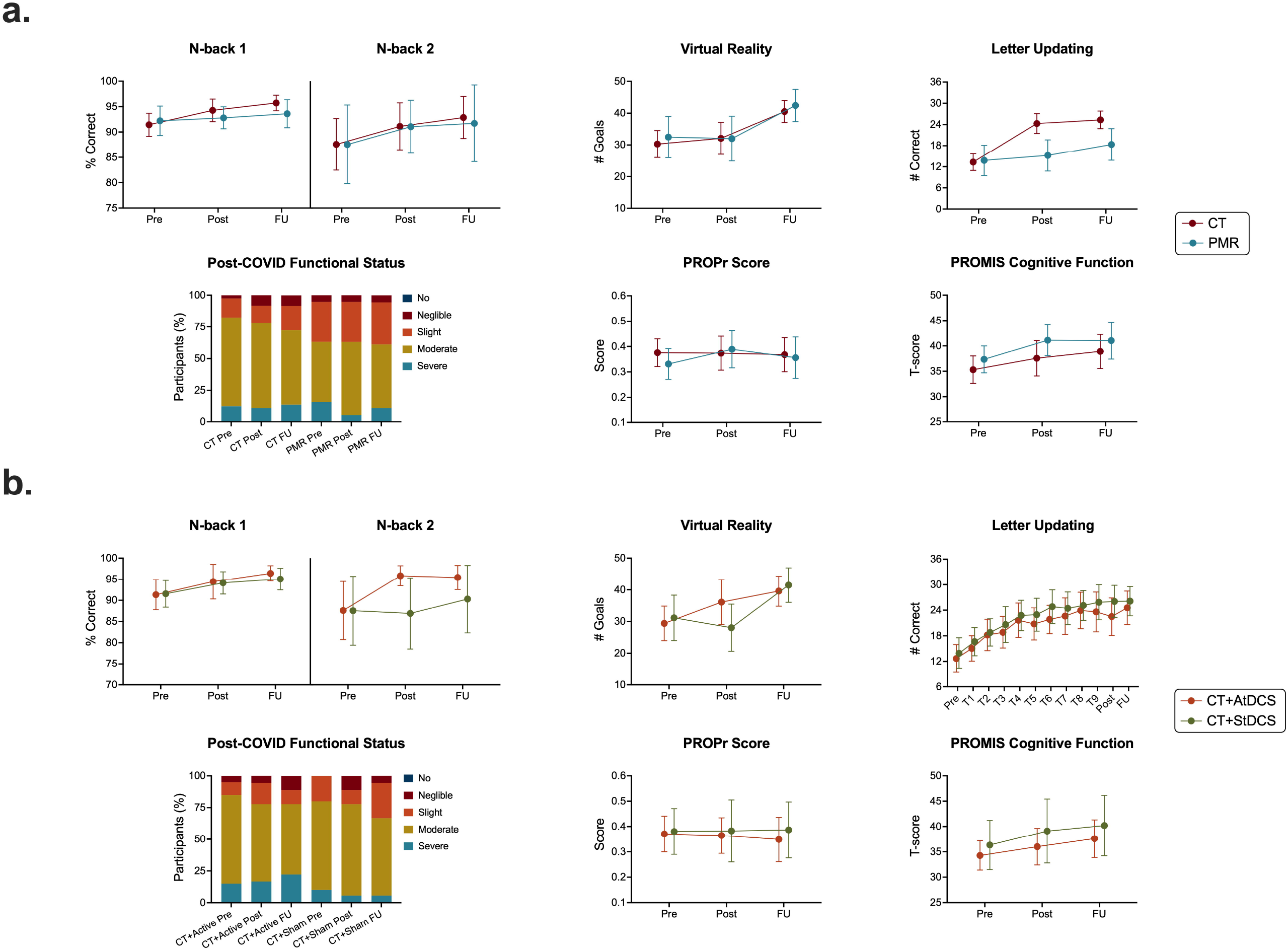
Raw cognitive and self-reported outcomes in the intention-to-treat population. **(a)** Cognitive training (*n* = 40) vs. progressive muscle relaxation (*n* = 20). **(b)** Cognitive training with active tDCS (*n* = 20) vs. cognitive training with sham tDCS (*n* = 20). Data are raw mean scores with 95% CI at pre-intervention, post-intervention, and 1-month follow-up. Stacked bar charts show the distribution of Post-COVID Functional Status categories at each timepoint. CT = cognitive training; CT+AtDCS = cognitive training with active transcranial direct current stimulation; CT+StDCS = cognitive training with sham transcranial direct current stimulation; PMR = progressive muscle relaxation; PROMIS-Cog = PROMIS Cognitive Function Scale; PROPr = PROMIS-Preference Score.

### 3.1. Primary outcome

The primary outcome, N-back task performance at post-intervention, did not differ significantly between CT and PMR groups (*β* = 1.59, 95% CI -1.30 to 4.48, *p* = 0.278) in either the 1-back (*β* = 2.52, 95% CI -1.27 to 6.31, *p* = 0.191) or 2-back condition (*β* = 0.66, 95% CI -3.12 to 4.44, *p* = 0.732).

### 3.2. Secondary outcomes

For secondary outcomes comparing CT and PMR, N-back performance at follow-up remained similar between groups in both task conditions. Performance on the virtual reality task did not differ substantially between groups at both assessment points. Letter updating performance was higher in CT than in PMR at both post-intervention (*β* = 9.45, 95% CI 6.35 to 12.55, *p* < 0.001) and follow-up (*β* = 7.83, 95% CI 4.79 to 10.88, *p* < 0.001). Functional status (Supplementary Table S4), health-related quality of life, and subjective cognitive function did not largely differ between groups at any timepoint.

For secondary outcomes comparing the CT groups, CT+AtDCS outperformed CT+StDCS in the 2-back condition at both post-intervention (*β* = 6.90, 95% CI 2.54 to 11.27, *p* = 0.002) and follow-up (*β* = 4.53, 95% CI 0.29 to 8.78, *p* = 0.037). No substantial differences emerged between groups in the 1-back condition at either timepoint. CT+AtDCS also outperformed CT+StDCS in the virtual reality task at post-intervention (*β* = 6.63, 95% CI 0.42 to 12.85, *p* = 0.037), but this effect was attenuated at follow-up. In the letter updating task, CT+StDCS yielded higher values than CT+AtDCS at post-intervention (*β* = -3.26, 95% CI -6.31 to -0.21, *p* = 0.036), with no larger difference at follow-up. Functional status (Supplementary Table S5), health-related quality of life, and subjective cognitive function did not largely differ between groups at any timepoint.

### 3.3. Safety outcomes

Overall, 52 AE occurred in 20 participants (incidence rate = 0.15, 95% CI 0.11 to 0.20). By treatment group, 17 events occurred in 7 CT+AtDCS participants (incidence rate = 0.16, 95% CI

0.09 to 0.24), 13 events in 6 CT+StDCS participants (incidence rate = 0.12, 95% CI 0.07 to 0.20), and 22 events in 7 PMR participants (incidence rate = 0.17, 95% CI 0.11 to 0.26). AE rates did not largely differ between CT and PMR (incidence rate ratio = 0.80, 95% CI 0.46 to 1.39) or between CT+AtDCS and CT+StDCS (incidence rate ratio = 1.31, 95% CI 0.64 to 2.75). No serious AE were reported and no participant discontinued the intervention due to AE. Detailed safety data are shown in Supplementary Table S6.

### 3.4. Blinding

Overall blinding was successful (James BI = 0.79, 95% CI 0.65 to 0.92). Detailed blinding data are presented in Supplementary Table S7.

## 4. Discussion

In this phase IIb PROBE trial, CT (pooled active and sham tDCS groups) did not improve untrained WM performance at post-intervention (primary outcome) or any secondary untrained cognitive measures compared with PMR in post-COVID-19 patients. However, CT combined with active tDCS enhanced untrained WM performance at post-intervention and 1-month follow-up and untrained visuospatial memory performance at post-intervention, compared with CT plus sham stimulation. Neither intervention substantially improved functional status, health-related quality of life, or subjective cognitive function. These findings provide the first controlled evidence jointly demonstrating that neuromodulation approaches may be necessary for cognitive transfer while highlighting the limited efficacy of standalone CT in post-COVID-19 populations.

CT did not relevantly improve untrained WM performance at post-intervention (primary outcome) compared with PMR. This finding contrasts with a previous trial that reported improvements in executive control following gamified CT in post-COVID-19 patients [11]. Several methodological differences may explain this discrepancy. First, the previous study assessed task-switching, an executive control component requiring cognitive flexibility and inhibitory control, rather than WM, which may respond differently to CT. Indeed, the trial failed to demonstrate improvement in its primary outcome, sustained attention, which reinforces the likely domain-specific nature of CT effects. Second, they used a gamified digital platform that likely provided a more multisensory, dynamic, and engaging environment than our laboratory-based paradigm. Third, their training was more intensive (≥ 5 sessions/week, 20-25 min/session, over six weeks) with flexible scheduling. Finally, their use of a passive control condition may have amplified apparent intervention effects.

Despite inducing immediate and sustained improvements on the trained task, CT showed no transfer to untrained WM or visuospatial memory measures at either timepoint. This pattern aligns with extensive research in healthy populations demonstrating that CT benefits are typically restricted to the trained tasks with minimal generalization to untrained measures [37– 39]. Such specificity likely reflects the acquisition of task-specific strategies and increased automatization rather than enhancement of transferable cognitive capacity [12]. These transfer limitations, along with the methodological factors mentioned earlier, suggest that effective CT for post-COVID-19 populations may require either protocols targeting broader executive domains with enhanced ecological validity and training intensity or adjunctive neuromodulation approaches that facilitate transfer effects.

Conversely, CT with active tDCS enhanced untrained WM performance at both timepoints compared with CT with sham stimulation. These findings contrast with a recent randomized controlled trial in post-COVID-19 patients, which reported no WM enhancement immediately after CT with tDCS [22]. This discrepancy may reflect methodological differences, including our WM-focused training vs. their multimodal intervention, our younger sample (18-60 years) vs. their broader age range (18-75 years), and our use of focal tDCS vs. their conventional approach. Our results partially align with a randomized trial in post-COVID-19 patients, which reported immediate and sustained WM gains after CT with active tDCS over the DLPFC [23]. However, that study assessed two different active montages without a sham condition, precluding conclusions about tDCS-specific efficacy. Our findings extend this preliminary evidence by demonstrating similar benefits in a rigorous sham-controlled design. Evidence from other clinical and healthy aging populations supports our findings. Meta-analyses across neurodegenerative and psychiatric disorders show superior WM outcomes for CT plus tDCS compared with CT alone [15]. Similarly, randomized controlled trials in neurodegenerative mild cognitive impairment (MCI) and healthy aging populations, using comparable protocols and the same stimulation target, have demonstrated immediate and sustained improvements in untrained WM performance [19,26]. Our results extend this evidence by showing that tDCS-augmented WM training can yield lasting improvements in WM in post-COVID-19 patients.

CT with active tDCS also improved untrained visuospatial memory performance at post-intervention compared with CT plus sham stimulation. Although no previous studies have combined WM training with DLPFC-tDCS to assess visuospatial outcomes, evidence from other clinical populations supports the plausibility of our finding. In patients with Alzheimer’s Disease dementia and MCI, single-session active tDCS over the right temporoparietal junction during visuospatial encoding enhanced subsequent recall compared with sham stimulation [40]. Similarly, three consecutive sessions of active tDCS over the same region during spatial memory training improved post-training performance in patients with MCI [41]. While these studies differ from ours in terms of training paradigms, dose, stimulation parameters, and populations, they converge in demonstrating immediate benefits to visuospatial memory from tDCS-augmented CT. Our findings extend this evidence by showing that DLPFC-targeted tDCS, even during non-visuospatial training, can facilitate cross-domain transfer to visuospatial memory in post-COVID-19 patients.

Neither CT nor CT with tDCS significantly improved post-COVID-19 functional status, health-related quality of life, or subjective cognitive function. This finding contrasts with a previous trial demonstrating functional and quality of life improvements following CT in post-COVID-19 patients [11]. Methodological differences, including training modality, higher dose intensity, and passive control condition, may explain this discrepancy. Similar quality of life benefits have been observed following CT with DLPFC-tDCS, although the absence of a sham condition precludes conclusions about stimulation-specific effects [23]. Our results are unsurprising given that this was a short-term, low-dose CT intervention targeting WM function. Transfer to self-report outcomes might require additional CT with tDCS boosting sessions or more intensive training protocols. Moreover, post-COVID-19 condition encompasses diverse physical, cognitive, and psychological symptoms that may require comprehensive rehabilitation rather than isolated cognitive interventions. Indeed, previous evidence suggests that both physical-mental health rehabilitation and rehabilitation-education programs, combined with motor cortex-targeted tDCS, can improve the quality of life in post-COVID-19 patients [21,42]. These findings suggest that multidisciplinary interventions addressing the multifactorial nature of post-COVID-19 symptoms may be necessary to maximize functional benefits.

Our findings have important implications for clinical practice and future research in post-COVID-19 populations. Notably, CT was insufficient to improve performance on untrained cognitive tasks, suggesting that current rehabilitation guidelines recommending standalone cognitive interventions for post-COVID-19 populations [43] may require refinement. Conversely, the selective improvements in WM and visuospatial memory performance observed with CT plus tDCS highlight the potential value of this combined approach. The intervention showed a favorable safety profile, with no serious AE and similar AE rates across groups, supporting the tolerability of tDCS as an adjunctive treatment. While these preliminary results are encouraging, their clinical significance requires validation through randomized controlled trials with larger sample sizes, multicenter designs, longer follow-up periods, and outcome measures that capture real-world functional improvements.

This study has several limitations. First, our sample was relatively small, recruited from a single site, predominantly female, and highly educated, which limits generalizability to broader post-COVID-19 populations. Future multicenter trials with larger, more diverse demographic samples should replicate our findings. Second, we did not assess relevant clinical variables such as vaccination status or viral variant that may influence cognitive outcomes and treatment response. Future studies should characterize these factors to identify potential moderators and predictors of treatment efficacy. Third, although cognitive benefits were observed at post-intervention and 1-month follow-up, this follow-up duration may be insufficient to determine the long-term stability of transfer effects. Longer follow-up periods, ideally spanning several months, are needed to assess the durability of effects and the need for booster or maintenance interventions. Fourth, the absence of neural or biomarker measures limits insight into the mechanisms underlying the observed behavioral effects and precludes the identification of predictors of treatment response. Future research should integrate multimodal approaches, including neuroimaging measures of structural and functional connectivity, electroencephalography, and biological markers relevant to post-COVID-19 pathophysiology, to elucidate mechanisms of action and treatment response.

In conclusion, this phase IIb trial provides the first controlled evidence in post-COVID-19 patients demonstrating that while CT alone does not improve untrained cognitive performance, CT combined with tDCS produces significant enhancements in transfer cognitive tasks. These findings highlight the potential value of combined neuromodulation approaches for cognitive rehabilitation in this population and should be validated in future phase III trials.

## Data Availability

The data supporting this study's findings are available from the corresponding author upon reasonable request.

## Acknowledgments

We thank all patients who participated in the study. We are also grateful to Robert Malinowski for providing technical support, Friederike Thams for her contributions to the study setup, Anke Steinmetz for assistance with patient recruitment, and Sein Schmidt for providing German-language assessment instruments.

## Funding

This project was funded by the German Research Foundation (Deutsche Forschungsgemeinschaft, DFG): Grant 497919823 to DA (AN 1103/4-1), Project number 539593253 to DA (AN 1103/6-1), Research Unit 5429/1 (467143400) to DA (AN 1103/5-1), AF (FL 379/34-1, FL 379/35-1) and MM (MM 3161/5-1, MM 3161/6-1) and CRC1315/B03 (327654276) to AF. CTLL is a PhD fellow funded by the German Academic Exchange Service (Deutscher Akademischer Austauschdienst, DAAD; scholarship: 91828451). The funders of the study had no role in study design, data collection, data analysis, data interpretation, or writing of the report.

## Notes

### Competing Interest Statement

The authors have declared no competing interest.

### Clinical Trial

NCT04944147

### Author Declarations

Ethics committee/IRB of University Medicine Greifswald gave ethical approval for this work

