## Supplementary_material for "Transcranial direct current stimulation-augmented cognitive training for post-COVID-19 cognition: A phase IIb randomized controlled trial"

### 1. Supplementary tables

#### Table S1. Cognitive baseline characteristics

|  | Overall  (*n* = 60) | CT  (*n* = 40) | PMR  (*n* = 20) | CT+AtDCS  (*n* = 20) | CT+StDCS  (*n* = 20) |
| --- | --- | --- | --- | --- | --- |
| **VLMT** |  |  |  |  |  |
| Immediate recall | 13.1 (2.0) | 12.9 (2.1) | 13.4 (1.5) | 13.1 (2.3) | 12.8 (2.0) |
| Delayed recall | 11.9 (3.2) | 11.7 (3.6) | 12.3 (2.4) | 11.8 (3.9) | 11.5 (3.3) |
| Learning | 55.2 (10.3) | 54.2 (11.0) | 57.1 (8.5) | 54.7 (11.7) | 53.6 (10.6) |
| Recognition | 14.2 (1.2) | 14.2 (1.3) | 14.2 (1.2) | 14.2 (1.5) | 14.3 (1.1) |
| **ROCF** |  |  |  |  |  |
| Copy | 35.5 (1.1) | 35.4 (1.2) | 35.6 (0.8) | 35.7 (0.7) | 35.2 (1.5) |
| Immediate retrieval | 26.5 (5.3) | 26.2 (5.4) | 27.3 (5.3) | 26.0 (5.3) | 26.3 (5.6) |
| Delayed retrieval | 26.0 (5.4) | 25.5 (5.7) | 27.1 (4.8) | 25.5 (5.9) | 25.5 (5.6) |
| **Digit span** |  |  |  |  |  |
| Forward | 6.9 (1.8) | 6.8 (1.9) | 7.0 (1.6) | 6.6 (1.8) | 7.2 (2.0) |
| Backward | 6.1 (1.7) | 6.2 (1.9) | 5.9 (1.3) | 6.3 (1.4) | 6.0 (2.3) |
| **TMT** |  |  |  |  |  |
| Part-A | 31.8 (13.4) | 34.4 (15.0) | 26.6 (7.2) | 34.7 (14.3) | 34.1 (16.1) |
| Part-B | 67.1 (22.4) | 69.1 (24.4) | 62.9 (17.6) | 69.0 (21.7) | 69.2 (27.5) |
| Stroop interference | 92.2 (31.6) | 93.9 (30.6) | 88.8 (34.0) | 94.1 (31.2) | 93.7 (30.8) |
| Phonological fluency | 12.9 (4.0) | 12.4 (3.9) | 13.8 (4.2) | 12.3 (3.5) | 12.6 (4.4) |
| Semantic fluency | 24.1 (5.8) | 23.2 (5.5) | 25.9 (6.1) | 23.2 (5.9) | 23.3 (5.3) |

Data are mean (*SD*). CT = cognitive training; CT+AtDCS = cognitive training with active transcranial direct current stimulation; CT+StDCS = cognitive training with sham transcranial direct current stimulation; PMR = progressive muscle relaxation; ROCF = Rey-Osterrieth Complex Figure; TMT = Trail Making Test; VLMT = Verbal Learning Memory Test.

#### Table S2. Raw outcomes in cognitive training and progressive muscle relaxation groups

|  | Pre-intervention | | Post-intervention | | 1-month follow-up | |
| --- | --- | --- | --- | --- | --- | --- |
|  | **CT**  (*n* = 40) | **PMR**  (*n* = 20) | **CT**  (*n* = 40) | **PMR**  (*n* = 20) | **CT**  (*n* = 40) | **PMR**  (*n* = 20) |
| N-back 1† | 91.4 (7.1) | 92.2 (6.2) | 94.3 (6.2) | 92.8 (4.5) | 95.7 (4.4) | 93.6 (5.9) |
| N-back 2† | 87.6 (15.9) | 87.6 (16.6) | 91.1 (13.3) | 91.1 (10.8) | 92.9 (12.2) | 91.7 (16.1) |
| Virtual Reality | 30.3 (13.0) | 32.5 (13.5) | 32.1 (14.5) | 32.0 (14.2) | 40.5 (9.8) | 42.4 (10.1) |
| Letter Updating | 13.3 (7.2) | 13.8 (9.2) | 24.3 (8.3) | 15.2 (9.2) | 25.3 (7.4) | 18.4 (9.6) |
| PCFS |  |  |  |  |  |  |
| Negligible | 1 (2.5%) | 1 (5.3%) | 3 (8.3%) | 1 (5.3%) | 3 (8.3%) | 1 (5.6%) |
| Slight | 6 (15.0%) | 6 (31.6%) | 5 (13.9%) | 6 (31.6%) | 7 (19.4%) | 6 (33.3%) |
| Moderate | 28 (70.0%) | 9 (47.4%) | 24 (66.7%) | 11 (57.9%) | 21 (58.3%) | 9 (50.0%) |
| Severe | 5 (12.5%) | 3 (15.8%) | 4 (11.1%) | 1 (5.3%) | 5 (13.9%) | 2 (11.1%) |
| PROPr Score | 0.4 (0.2) | 0.3 (0.1) | 0.4 (0.2) | 0.4 (0.2) | 0.4 (0.2) | 0.4 (0.2) |
| PROMIS-Cog | 35.3 (8.5) | 37.4 (5.6) | 37.6 (10.3) | 41.1 (6.3) | 38.9 (9.9) | 41.0 (7.8) |

Data are mean (*SD*) or *n* (%). †Primary outcome. CT = cognitive training; PCFS = Post-COVID Functional Status Scale; PMR = progressive muscle relaxation; PROMIS-Cog = PROMIS Cognitive Function Scale; PROPr = PROMIS-Preference Score.

#### Table S3. Raw outcomes in cognitive training with active and sham tDCS groups

|  | Pre-intervention | | Post-intervention | | 1-month follow-up | |
| --- | --- | --- | --- | --- | --- | --- |
|  | **CT+AtDCS**  (*n* = 20) | **CT+StDCS**  (*n* = 20) | **CT+AtDCS**  (*n* = 20) | **CT+StDCS**  (*n* = 20) | **CT+AtDCS**  (*n* = 20) | **CT+StDCS**  (*n* = 20) |
| N-back 1 | 91.3 (7.6) | 91.6 (6.7) | 94.4 (7.4) | 94.1 (5.3) | 96.4 (3.5) | 95.1 (5.2) |
| N-back 2 | 87.6 (14.7) | 87.6 (17.4) | 95.8 (4.4) | 86.9 (16.9) | 95.4 (5.8) | 90.3 (16.0) |
| Virtual Reality | 29.4 (11.3) | 31.2 (14.8) | 36.1 (13.8) | 28.1 (14.5) | 39.6 (9.2) | 41.5 (10.6) |
| Letter Updating | 12.7 (6.8) | 13.9 (7.8) | 22.5 (8.8) | 26.1 (7.6) | 24.6 (7.9) | 26.1 (6.9) |
| PCFS |  |  |  |  |  |  |
| Negligible | 1 (5.0%) | 0 (0.0%) | 1 (5.6%) | 2 (11.1%) | 2 (11.1%) | 1 (5.6%) |
| Slight | 2 (10.0%) | 4 (20.0%) | 3 (16.7%) | 2 (11.1%) | 2 (11.1%) | 5 (27.8%) |
| Moderate | 14 (70.0%) | 14 (70.0%) | 11 (61.1%) | 13 (72.2%) | 10 (55.6%) | 11 (61.1%) |
| Severe | 3 (15.0%) | 2 (10.0%) | 3 (16.7%) | 1 (5.6%) | 4 (22.2%) | 1 (5.6%) |
| PROPr Score | 0.4 (0.1) | 0.4 (0.2) | 0.4 (0.1) | 0.4 (0.2) | 0.3 (0.2) | 0.4 (0.2) |
| PROMIS-Cog | 34.3 (6.2) | 36.3 (10.4) | 36.0 (7.2) | 39.1 (12.7) | 37.6 (7.5) | 40.2 (12.0) |

Data are mean (*SD*) or *n* (%). CT+AtDCS = cognitive training with active transcranial direct current stimulation. CT+StDCS = cognitive training with sham transcranial direct current stimulation; PCFS = Post-COVID Functional Status Scale; PROMIS-Cog = PROMIS Cognitive Function Scale; PROPr = PROMIS-Preference Score.

#### Table S4. Post-COVID functional status outcomes in cognitive training and progressive muscle relaxation groups

|  | CT  Adjusted probability (95% CI) | PMR  Adjusted probability (95% CI) | Group difference  OR (95% CI) | *p*-value |
| --- | --- | --- | --- | --- |
| Intention-to-treat analysis ^a^ |  |  |  |  |
| Post-intervention | 80.1% (59.2–91.8%) | 76.3% (44.9–92.7%) | 1.25 (0.28–5.52) | 0.765 |
| 1-month follow-up | 73.2% (50.1–88.1%) | 77.5% (40.6–94.5%) | 0.79 (0.13–4.70) | 0.795 |
| Per-protocol analysis ^b^ |  |  |  |  |
| Post-intervention | 75.0% (47.3–91.0%) | 69.3% (27.3–93.1%) | 1.33 (0.25–7.17) | 0.734 |
| 1-month follow-up | 64.4% (32.5–87.1%) | 67.9% (22.8–93.8%) | 0.85 (0.12–6.27) | 0.874 |

Data are probabilities (95% CI) of moderate-to-severe functional limitations (PCFS scores ≥3) and odds ratios comparing cognitive training and progressive muscle relaxation groups. ^a^ CT (*n* = 40); PMR (*n* = 20). ^b^ CT (*n* = 24); PMR (*n* = 14). PCFS scores were dichotomized into negligible-to-slight (grades 1-2) vs. moderate-to-severe (grades 3-4) functional limitations due to sparse cell frequencies. CT = cognitive training; PCFS = Post-COVID Functional Status Scale; PMR = progressive muscle relaxation.

#### Table S5. Post-COVID functional status outcomes in cognitive training with active and sham tDCS groups

|  | CT+AtDCS  Adjusted probability (95% CI) | CT+StDCS  Adjusted probability (95% CI) | Group difference  OR (95% CI) | *p*-value |
| --- | --- | --- | --- | --- |
| Intention-to-treat analysis ^a^ |  |  |  |  |
| Post-intervention | 87.7% (54.6–97.7%) | 84.2% (54.7–95.9%) | 1.34 (0.20–8.80) | 0.755 |
| 1-month follow-up | 81.2% (46.8–95.5%) | 76.3% (38.8–94.2%) | 1.34 (0.20–8.80) | 0.755 |
| Per-protocol analysis ^b^ |  |  |  |  |
| Post-intervention | 32.5% (5.4–80.2%) | 19.8% (1.8–76.8%) | 1.94 (0.23–16.33) | 0.540 |
| 1-month follow-up | 19.5% (2.7–68.0%) | 11.1% (0.6–70.3%) | 1.94 (0.23–16.33) | 0.540 |

Data are probabilities (95% CI) of moderate-to-severe functional limitations (PCFS scores ≥3) and odds ratios comparing cognitive training with active tDCS vs. cognitive training with sham tDCS groups. ^a^ CT+AtDCS (*n* = 20); CT+StDCS (*n* = 20). ^b^ CT+AtDCS (*n* = 12); CT+StDCS (*n* = 12). PCFS scores were dichotomized into negligible-to-slight (grades 1-2) vs. moderate-to-severe (grades 3-4) functional limitations due to sparse cell frequencies. CT+AtDCS = cognitive training with active transcranial direct current stimulation. CT+StDCS = cognitive training with sham transcranial direct current stimulation; PCFS = Post-COVID Functional Status Scale.

#### Table S6. Adverse events by treatment group and sensation type

|  | **Overall**  (*n* = 38) | | **CT+AtDCS**  (*n* = 12) | | **CT+StDCS**  (*n* = 12) | | **PMR**  (*n* = 14) | | **CT**  **vs.**  **PMR** | **CT+AtDCS**  **vs.**  **CT+StDCS** |
| --- | --- | --- | --- | --- | --- | --- | --- | --- | --- | --- |
|  | Participants/ Events | IR (95% CI) | Participants/ Events | IR (95% CI) | Participants/ Events | IR (95% CI) | Participants/ Events | IR (95% CI) | IRR (95% CI) | IRR (95% CI) |
| **All events** | 20/52 | 0.15  (0.11–0.20) | 7/17 | 0.16 (0.09–0.24) | 6/13 | 0.12 (0.07–0.20) | 7/22 | 0.17 (0.11–0.26) | 0.80 (0.46–1.39) | 1.31 (0.64–2.75) |
| **Itching** | 6/8 | 0.02  (0.01–0.04) | 1/1 | 0.01 (0–0.04) | 2/3 | 0.03 (0.01–0.07) | 3/4 | 0.03 (0.01–0.07) | 0.58 (0.14–2.47) | 0.33 (0.02–2.60) |
| **Pain** | 5/8 | 0.02  (0.01–0.04) | 2/3 | 0.03 (0.01–0.07) | 1/1 | 0.01 (0–0.04) | 2/4 | 0.03 (0.01–0.07) | 0.58 (0.14–2.47) | 3.00 (0.38–60.65) |
| **Burning** | 9/12 | 0.04  (0.02–0.06) | 3/4 | 0.04 (0.01–0.09) | 2/2 | 0.02 (0–0.06) | 4/6 | 0.05 (0.02–0.10) | 0.58 (0.18–1.86) | 2.00 (0.39–14.43) |
| **Heat** | 4/7 | 0.02  (0.01–0.04) | 2/3 | 0.03 (0.01–0.07) | 1/2 | 0.02 (0.00–0.06) | 1/2 | 0.02 (0–0.05) | 1.46 (0.31–10.18) | 1.50 (0.25–11.39) |
| **Metallic taste** | 0/0 | - | 0/0 | - | 0/0 | - | 0/0 | - | - | - |
| **Fatigue** | 6/10 | 0.03  (0.01–0.05) | 2/4 | 0.04 (0.01–0.09) | 2/3 | 0.03 (0.01–0.07) | 2/3 | 0.02 (0.01–0.06) | 1.36 (0.38–6.32) | 1.33 (0.29–6.77) |
| **Other sensations** | 4/7 | 0.02 (0.01–0.04) | 1/2 | 0.02 (0–0.06) | 1/2 | 0.02 (0–0.06) | 2/3 | 0.02 (0.01–0.06) | 0.78 (0.17–3.95) | 1.00 (0.12–8.33) |

Data are the number of participants experiencing events/total number of events, with incidence rates (events per person-day) and 95% CI in the safety analysis set. Incidence rate ratios compare cognitive training vs. progressive muscle relaxation and cognitive training with active vs. cognitive training with sham transcranial direct current stimulation. CT+AtDCS = cognitive training with active transcranial direct current stimulation; CT+StDCS = cognitive training with sham transcranial direct current stimulation; IR = incidence rate; IRR = incidence rate ratio; PMR = progressive muscle relaxation.

#### Table S7. Blinding effectiveness in cognitive training groups

|  | Overall  (*n* = 28) | CT+AtDCS  (*n* = 14) | CT+StDCS  (*n* = 14) |
| --- | --- | --- | --- |
| Treatment assignment guess |  |  |  |
| Active tDCS | 12 (42.9%) | 7 (50.0%) | 5 (35.7%) |
| Sham tDCS | 4 (14.3%) | 1 (7.1%) | 3 (21.4%) |
| Don't know | 12 (42.9%) | 6 (42.9%) | 6 (42.9%) |

Data are *n* (%) in the intention-to-treat population. Of 40 participants randomized to CT groups, 28 (70%) provided valid blinding assessment data. Missing data included five participants who completed visit 10 but did not respond to the blinding question (CT+AtDCS: *n* = 3; CT+StDCS: *n* = 2) and seven participants who did not attend visit 10 (CT+AtDCS: *n* = 3; CT+StDCS: *n* = 4). CT+AtDCS = cognitive training with active transcranial direct current stimulation; CT+StDCS = cognitive training with sham transcranial direct current stimulation.

### 2. Supplementary figures

#### Figure S1. Primary and secondary outcomes in the per-protocol population

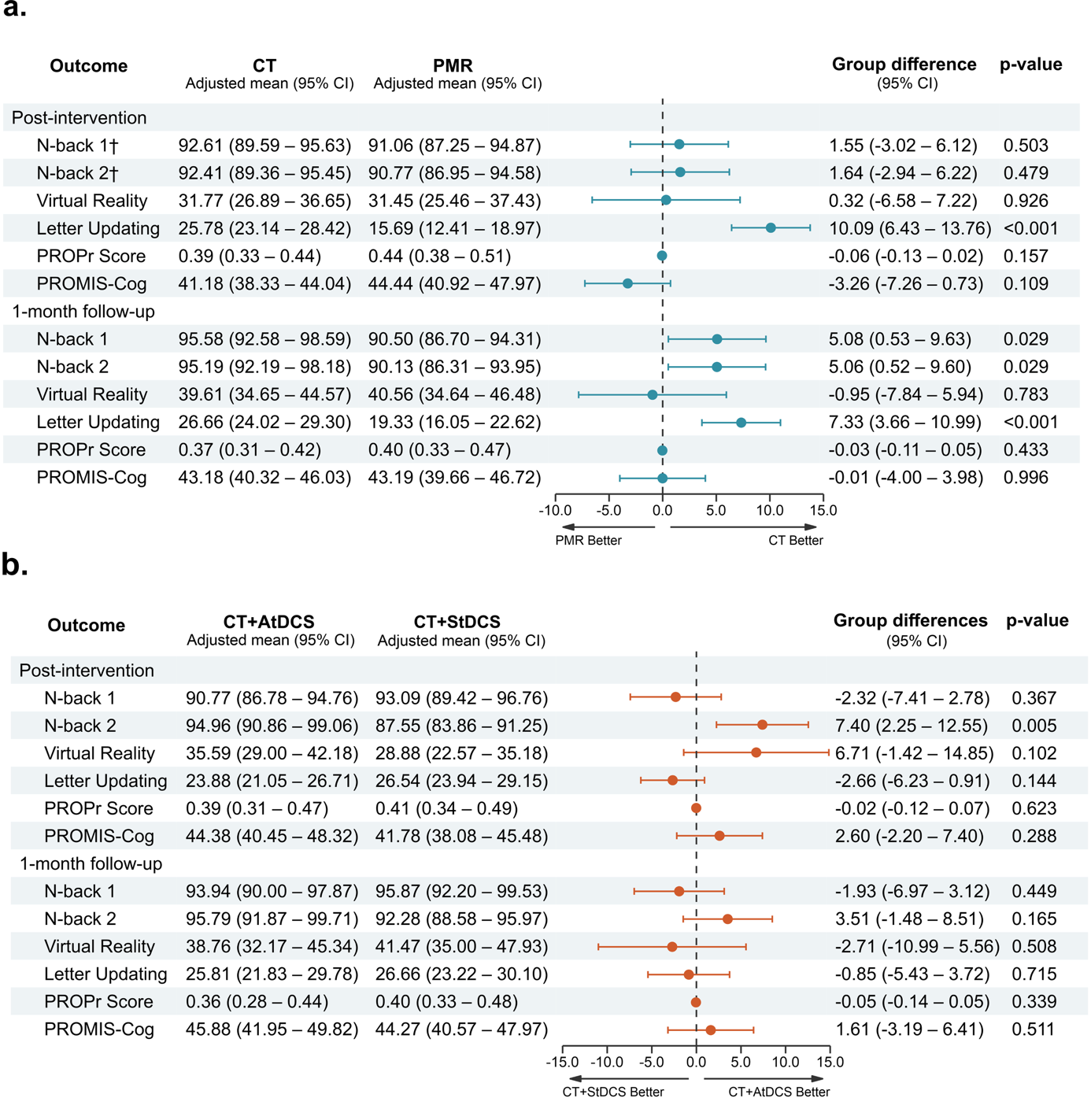

**Figure S1. Results for primary and secondary outcomes in the per-protocol population. (a)** Cognitive training (*n* = 24) vs. progressive muscle relaxation (*n* = 14). **(b)** Cognitive training with active tDCS (*n* = 12) vs. cognitive training with sham tDCS (*n* = 12). Forest plots show adjusted between-group mean differences with 95% CI at post-intervention and 1-month follow-up, derived from linear mixed-effects models using raw data. Positive values favor cognitive training in panel (a) and cognitive training with active tDCS in panel (b). ^†^Primary outcome. CT = cognitive training; CT+AtDCS = cognitive training with active transcranial direct current stimulation; CT+StDCS = cognitive training with sham transcranial direct current stimulation; PMR = progressive muscle relaxation; PROMIS-Cog = PROMIS Cognitive Function Scale; PROPr = PROMIS-Preference Score.
